# Day 3 parasitemia and *Plasmodium falciparum Kelch 13* mutations among uncomplicated malaria patients treated with artemether-lumefantrine in Adjumani district, Uganda

**DOI:** 10.1101/2024.04.26.24306433

**Authors:** Martin Kamilo Angwe, Norah Mwebaza, Sam Lubwama Nsobya, Patrick Vudriko, Savior Dralabu, Denis Omali, Maria Agnes Tumwebaze, Moses Ocan

## Abstract

Artemisinin resistance threatens malaria control and elimination efforts globally. Recent studies have reported the emergence of *Plasmodium falciparum* parasites tolerant to artemisinin agents in sub-Saharan Africa, including Uganda. The current study assessed the day 3 parasite clearance and its correlation with *P. falciparum K13* propeller gene (*pfkelch13*) mutations in *P. falciparum* parasites isolated from patients with uncomplicated malaria under artemether-lumefantrine (AL) treatment. This study enrolled 100 *P. falciparum-*positive patients to whom AL was prescribed between 09/September/2022 and 06/November/2022. Blood samples were collected in EDTA tubes before treatment initiation (day 0) and on day 3. Parasitemia was assessed by microscopy from blood smears and quantitative polymerase chain reaction (qPCR) from the DNA extracted. The day 0 parasite *K13* gene was sequenced using Sanger sequencing. Sequence data were analysed using MEGA *version* 11 software. The data were analysed using STATA *version* 15, and the Mann‒Whitney U test was used to compare PCR parasite clearance on day 3 using the comparative CT value method and *pfkelch13* mutations.

The prevalence of day 3 parasitaemia was 24% (24/100) by microscopy and 63% (63/100) by qPCR from the AL-treated patients. *P. falciparum K13*-propeller gene polymorphism was detected in 18.8% (15/80) of the day 0 DNA samples. The *K13* mutations found were C469Y, 12.5% (10/80); A675V, 2.5% (2/80); A569S, 1.25%, (1/80), A578S, 1.25%, (1/80) and; F491S, 1.25%, (1/80) a new allele not reported anywhere. The C469Y mutation, compared to the wild-type, was associated with delayed parasite clearance *p*=0.0278, Hodges-Lehmann estimation 3.2108 on the log scale, (95%CI 1.7076, 4.4730).

There was a high prevalence of day 3 *P. falciparum* among malaria patients treated using artemether-lumefantrine. We conclude that the *K13* mutation associated with artemisinin resistance by *P. falciparum* is present in Adjumani district, Uganda. This necessitates regular surveillance of the effectiveness and efficacy of artemether-lumefantrine in the country.

## Introduction

Malaria continues to pose a substantial global health challenge, with sub-Saharan Africa (SSA) bearing the highest burden of the disease’s impact. In 2022, the World Health Organization (WHO) reported 249 million malaria cases worldwide, an increase of at least 5 million cases compared to the previous year, with the WHO African region accounting for 94% of the cases (1,2). Uganda contributes at least 5% of the worldwide malaria cases, most of which occur in rural areas, especially in Northern and Eastern Uganda. There have been extensive control and prevention interventions, such as the distribution of insecticide-treated bed nets (ITNs) covering at least 82% of households in Uganda, indoor residual spraying (IRS) in some regions being scaled up, and antimalarial medications. Despite these interventions, the area has witnessed stagnation in malaria elimination efforts and an increase in malaria cases in Northern Uganda, highlighting the persistent challenge of malaria, especially in resource-limited settings (3).

The current malaria treatment approach for uncomplicated *Plasmodium falciparum* infection involves using artemisinin-based combination therapies (ACTs). Uganda adopted artemether-lumefantrine (AL) as a first-line therapy for uncomplicated *P. falciparum* malaria, which has been crucial in reducing malaria mortalities for almost two decades (1,4–6). These therapies combine the short-acting artemisinin derivative (artemether) with another long-acting antimalarial drug (lumefantrine), enhancing efficacy and reducing the risk of resistance development. In addition to case management by the ACTs, control measures such as ITNs, IRS, and malaria chemoprevention have been integral in reducing malaria transmission (7–12).

Despite these efforts, several challenges persist in the fight against malaria. Inadequate supply of antimalarial medicines, especially in remote areas, often results in stockouts of ACTs, the cornerstone of malaria treatment (13). Moreover, the proliferation of substandard antimalarial agents and self-medication pose a significant public health risk, undermining treatment effectiveness and contributing to artemisinin resistance development (14,15). Artemisinin resistance, characterised by *Pfkelch*13 (K13) gene mutations (16) and delayed parasite clearance, has emerged as a serious concern, and molecular markers of artemisinin resistance have been identified in many SSA countries (17,18). Recent studies in Uganda reported the emergence of artemisinin resistance in the Eastern and Northern regions (19–21), a threat that is becoming evident to the effectiveness of artemisinin combination therapy.

The widespread development of artemisinin resistance could reverse the gains made in malaria control and elimination, potentially leading to an increase in malaria morbidity and mortality. This may lead to a similar catastrophic event previously observed when chloroquine was withdrawn from malaria treatment due to widespread resistance (22,23). In Uganda, where healthcare systems are often overburdened with limited access to quality healthcare (24,25), the spread of artemisinin resistance could undermine decades of progress and investment in malaria control, and this could be worsen by the lack of effective alternatives to artemisinin antimalarial agents in malaria treatment.

This study assessed the prevalence of day 3 *P. falciparum* in patients with uncomplicated malaria treated with AL and the prevalence of K13-propeller gene mutations associated with artemisinin resistance. The study’s findings provide context-specific evidence on artemisinin resistance among *P. falciparum* parasites in patients treated using artemether-lumefantrine in the West Nile region, Uganda.

## Materials and Methods

### Ethical statement

The protocol was reviewed, and ethical approval was obtained from the School of Biomedical Sciences Research and Ethics Committee at the College of Health Sciences, Makerere University (SBS-2022-157). Administrative clearance was obtained from Adjumani district authorities. Written informed consent was obtained from all participants before data collection, parental consent was obtained from participants aged 5-7 years, assent and parental consent were obtained from participants aged 8-17 years, and participants 18 years old and above consented to the study.

### Study area

The study was conducted in the Adjumani district located in the West Nile region, northwest of Uganda. The region neighbours the Democratic Republic of Congo and South Sudan. The district is among the highest malaria endemic districts in the country, with malaria transmission throughout the year and a prevalence reported above 50% by microscopy during the country’s malaria indicator survey (26).

### Study design

This was a prospective longitudinal study which recruited malaria patients between 09/September/2022 and 06/November/2022 receiving artemether-lumefantrine (AL) 20 mg artemether/120 mg lumefantrine tablets, 3-day six-dose regimen prescribed according to patient weight for treatment of uncomplicated malaria at Adjumani District Hospital. Blood samples were collected from the patient on day 0 (before drug intake) and day 3 (after treatment). The study enrolled 100 purposively selected uncomplicated malaria patients aged five years and above with confirmed *P. falciparum* infection. A study purpose non-validated questionnaire was used to collect information on the socio-demographic characteristics of the patients, including age, gender, and previous history of malaria treatment. The study collected both patient socio-demographic and laboratory data. *Plasmodium falciparum* parasite infections among symptomatic malaria patients were determined using microscopy and qPCR from the College of Veterinary Medicine Animal Resources and Biosecurity, Pharmacology/Research Center for Tropical Diseases and Vector Control, and Central Diagnostic Laboratory.

### Blood sample collection

Venous blood (2-3ml) was collected in an EDTA tube. Two blood samples were collected from each patient; the first was on day 0, before initiation of treatment, and the second was collected on day 3, after completion of artemether-lumefantrine therapy.

### Microscopy

On days 0 and 3, thick and thin smears were prepared for detection and speciation of *Plasmodium falciparum* infection, respectively. The blood smears were stained with 10% Giemsa stain for 30 minutes, then examined using an Olympus microscope under 100X magnification with oil immersion to identify the parasite and determine the parasite density. Two independent microscopists/laboratory technologists examined the slides for *P. falciparum* confirmation and enumeration.

### DNA extraction

The parasite DNA was extracted using a Qiagen DNA mini kit (QIAamp DNA Blood Mini Kit-250, Germany), following the manufacturer’s instructions. Elution was performed in 50 µL elution buffer, and the DNA concentration was determined (25-40 ng/µl) using NanoDrop (NanoDrope Lite Plus, thermo scientific) before PCR amplification.

### Plasmodium falciparum qPCR

Quantitative PCR (qPCR) was performed using a real-time PCR system (QuantStudio^TM^5) to detect and quantify the *Plasmodium falciparum* in duplicate. Primers and probes were based on the 18S rRNA sequence (27) synthesised from Inqaba Biotec Laboratory, South Africa. The amplification followed a method in a previous study (28). The CT values were recorded and used to calculate parasite clearance through fold change, 2^−ΔΔCT^ by a comparative CT value method (29). The method considered two data points of CT values at day 0 and day 3 for individuals that remained positive on day 3.

Specific primers were used for the qPCR (Table 1); the probe was labelled with 5′FAM (6-carboxyfluorescein) and BHQ-1^TM^ (Black Hole Quencher) as the reporter and quencher, respectively. 20 µl reaction volume was prepared, composed of 0.8 µl of each forward and reverse primer, 10 µl of qPCR master mix (Luna Universal Probe qPCR Master Mix NEB, New England BioLabs Inc), 0.4 µl of the probes, 20 ng of the DNA template and nuclease- free water (variable). The PCR conditions involved one cycle at 94°C for 5 min of initial denaturation, followed by 40 cycles of denaturation at 94°C for 30 s, annealing at 54°C for 90 s and extension at 68°C for 90 s, and a final extension at 68°C for 90 s.

**Table 1:**
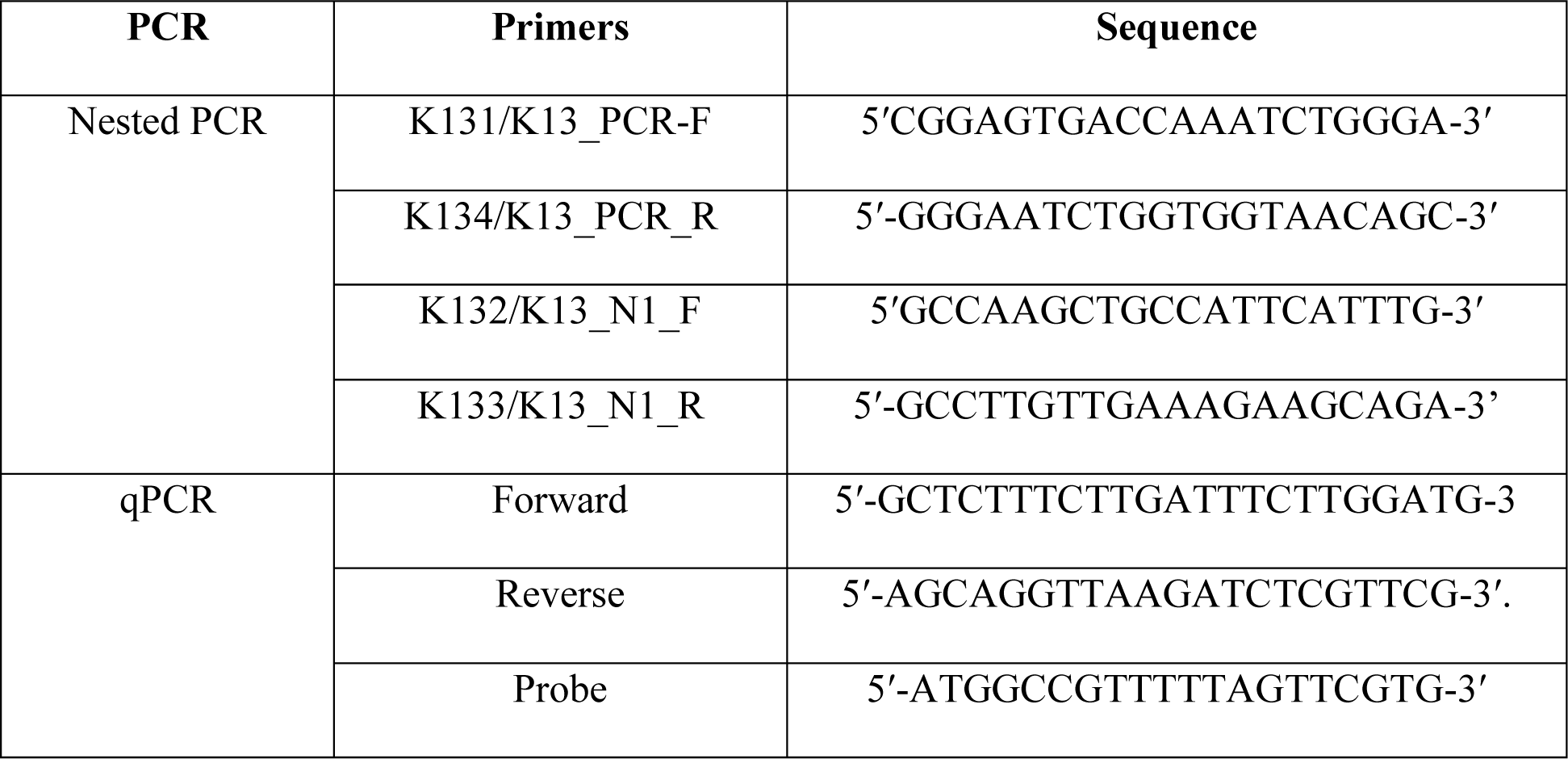
Primer sequences used for nested PCR and qPCR.

### P. falciparum K13 allele

Conventional nested PCR was performed (SimpliAmp Thermal Cycler, applied biosystem) to amplify approximately 800 bp fragment that extends from nucleotides position 1329-to-2178 (codons 445-to-727) within the propeller domain of the K13 gene using the specific primers (16) Table 1.

### *P. falciparum K13* gene amplification

The first round PCR was performed in a 25 µl reaction volume of 12.5 µl OneTaq 2X Master Mix (NEB, New England BioLabs Inc), 1 µl of each forward and reverse primer and 2.5 ng of the DNA template and nuclease-free water (variable). The PCR was run with the following conditions: 94°C for 5 m initial denaturation, 40 cycles at 94°C for 30 s, annealing at 54°C for 90 s, and extension at 68°C for 90 s. The final extension was performed at 68°C for 10 minutes, then kept at 4°C, which was set to infinite. The amplicon from the first round of PCR was used as a template for the second round. This was run in a 25 µL reaction mixture volume; it involved the same master mix preparation as the first run, but the second primer sets were used with 1 µl of the first PCR product/amplicon. The PCR product was visualised using gel electrophoresis run at 100 V for 30 minutes in 1.5% agarose stained with 0.05 µg/mL Acridine orange (SafeView Classic).

### Amplicon preparation and cleaning

The nested PCR products were cleaned using ExoSAP. The Exo/SAP master mix was prepared by adding 50 µL of exonuclease I (Catalogue No. NEB M0293) 20 U/µl and 200 µl of shrimp alkaline phosphatase (Catalogue No. NEB M0371) 1 U/µl to a 0.6 ml microcentrifuge tube. Then, 2.5 µl of the ExoSAP mixture was mixed with 10 µL of the amplicon/PCR product and incubated at 37°C for 15 min; the reaction was stopped by heating the mixture at 80°C for 15 min.

### Plasmodium falciparum K13 Sequencing

The nested PCR products were sequenced using the Nimagen BrilliantDye^TM^ Terminator Cycle Sequencing kit V3.1, BRD3-100/1000, according to the manufacturer’s instructions at the Inqaba Biotec laboratory in South Africa, a commercial sequencing company. The labelled product was then cleaned with a ZR-96 DNA sequencing clean-up Kit (Catalogue No. D4053). The cleaned product was injected into an Applied Biosystems ABI 3730XL Genetic Analyser with a 50 cm array using POP7. Sequence chromatogram analysis was performed using FinchTV analysis software.

### Data management and analysis

The K13 sequence qualities were assessed, and then BLAST against the NCBI database to confirm the identity of the gene sequence. After exporting the sequence into the Bioedit sequence alignment editor, manual editing was performed, and the sequence was exported into MEGA11 software for polymorphism detection using the sequence PF3D7_1343700 from the NCBI database as the reference sequence. Sequence alignment was performed using the ClustalW algorithm, and a phylogenetic tree was drawn using MEGA11 Tree Explorer to identify divergence in the *P. falciparum K13* genes and identify the mutations. Data were entered in Excel and analysed in STATA version 17; CT value fold change from the qPCR data was log-transformed before correlation. The relationship between the CT value fold change related to parasite clearance and K13 mutation was determined through a nonparametric test (Mann-Whitney U test) with a 95% confidence interval and level of significance set at *p*<0.05.

## Results

### Characteristics of the study participants

The study included 100 symptomatic uncomplicated malaria patients, of which the majority, 65% (65/100), were females. The mean age of the study participants was 17±10 years, ranging from 5 to 46 years. Parasitemia/µl geometric mean by microscopy was 3868, IQR 11104 for all patients.

### Prevalence of *P. falciparum* on day three after AL treatment

*P. falciparum* parasites persisted in 24% (24/100; 95%CI: 16-33) of the participants screened using microscopy, whereas by PCR, the parasites were found in over half, 63% (63/100; 95%CI: 53-72) on day 3 following artemether-lumefantrine treatment (Table 2).

**Table 2:** Day 3 P. falciparum positivity distribution among the different age groups.

| Characteristics | Microscopy and PCR<br>positive, day 0 (N=100) | Day 3 positivity |  |
| --- | --- | --- | --- |
|  |  | Microscopy, n (%) | PCR n (%) |
| Age |  |  |  |
| 5-12 | 35 | 10 (28.57) | 23 (65.71) |
| 13-18 | 37 | 9 (24.32) | 24 (64.86) |
| 19 and above | 28 | 5 (17.86) | 16 (57.14) |
| Sex |  |  |  |
| Male n=35 | 35 | 7 (20.00) | 22 (62.86) |
| Female n=65 | 65 | 17 (26.15) | 41 (63.08) |
| Total Participants | 100 | 24 (24.00) | 63 (63.00) |

N=number of participants; n=number of participants who remained positive on day 3

### Prevalence of *Pfkelch13* Single nucleotide polymorphism (SNPs)

A total of 80 samples (amplicons) out of the 100 were successfully sequenced for K13 Single Nucleotide Polymorphism (SNP) identification. Of the 80 sequenced samples, 24/80 were the day 3 microscopy positive, and 58/80 were day 3 PCR positive samples; the 20/100 unsequenced samples contain 5/20 day 3 qPCR positive samples, but all microscopy negative (20/20). K13 SNPs were detected in 18.8% (15/80; 95%CI: 11.25-27.5) of the *P. falciparum* parasite samples sequenced (Table 3). All the K13 SNPs were nonsynonymous mutations. The most prevalent nonsynonymous K13 SNP was C469Y, at 12.5% (10/80; 95%CI: 6.25-20). Other K13 SNPs in the *P. falciparum* parasites were A675V (2.25%, 2/80; 95%CI: 0.0-6.25), A569S (1.25%, 1/80; 95%CI:0.0-3.75) and A578S (1.25%, 1/80; 95%CI: 0.0-3.75). One mutation not previously reported was detected in one sample where amino acid F was substituted by S at codon 491 in the K13 propeller domain, involving a nucleotide change from T to C at position 1472.

**Table 3:** Prevalence of K13 single-nucleotide polymorphisms in P. falciparum parasites among patients in Adjumani District.

| Mutations | Wild-type amino acid | Mutant amino acid | Codon position | Wild-type nucleotide | Mutant nucleotide | Mutation type | Nucleotide position | Prevalence (n=80) |
| --- | --- | --- | --- | --- | --- | --- | --- | --- |
| C469Y<sup>ab</sup> | Cysteine | Tyrosine | 469 | G | A | NS | 1406 | 12.50% (n=10) |
| A675V<sup>ab</sup> | Alanine | Valine | 675 | C | T | NS | 2024 | 2.50% (n=2) |
| A569S<sup>a</sup> | Alanine | Serine | 569 | G | T | NS | 1705 | 1.25% (n=1) |
| A578S<sup>a</sup> | Alanine | Serine | 578 | G | T | NS | 1732 | 1.25% (n=1) |
| F491S | Phenylalanine | Serine | 491 | T | C | NS | 1491 | 1.25% (n=1) |

NS=non synonymous mutation; n=number of samples sequenced; <sup>a</sup>Previously reported *K13* allele, <sup>ab</sup>Candidate for artemisinin resistance

### Relationship between day 3 *P. falciparum* clearance and *Pfkelch13* Single nucleotide polymorphism

Eight (8/10) patients with parasites with C469Y and one (1/1) patient with the A578S *P. falciparum K13* mutation remained positive on day three by PCR. By microscopy, 5/10 patients with *P.falciparum* having C469Y and 1/1 having A578S remained positive on day 3 (Table 4). A comparison of the parasite clearance between the wild-type, median parasite fold reduction 522, 7043 (median, IQR) and mutated K13 parasites showed significantly low parasite clearance in C469Y, median fold reduction 14, 792 (Median, IQR) and A578S, fold reduction 1.6. Among the day 3 qPCR-positive *P. falciparum* patients, those with the C469Y mutated *P. falciparum* demonstrated low parasite clearance compared to the wild-type *p*=0.0278, Hodges-Lehmann estimation 3.2108 on the log scale (95%CI 1.7076, 4.4730). Parasitemia/µl geometric mean for patients with wild-type *P. falciparum* 65/80 was 3795, IQR 10589, while that for the patients with C469Y *P. falciparum* (10/80) was 3373, IQR 26099 for the sequenced samples.

**Table 4:**
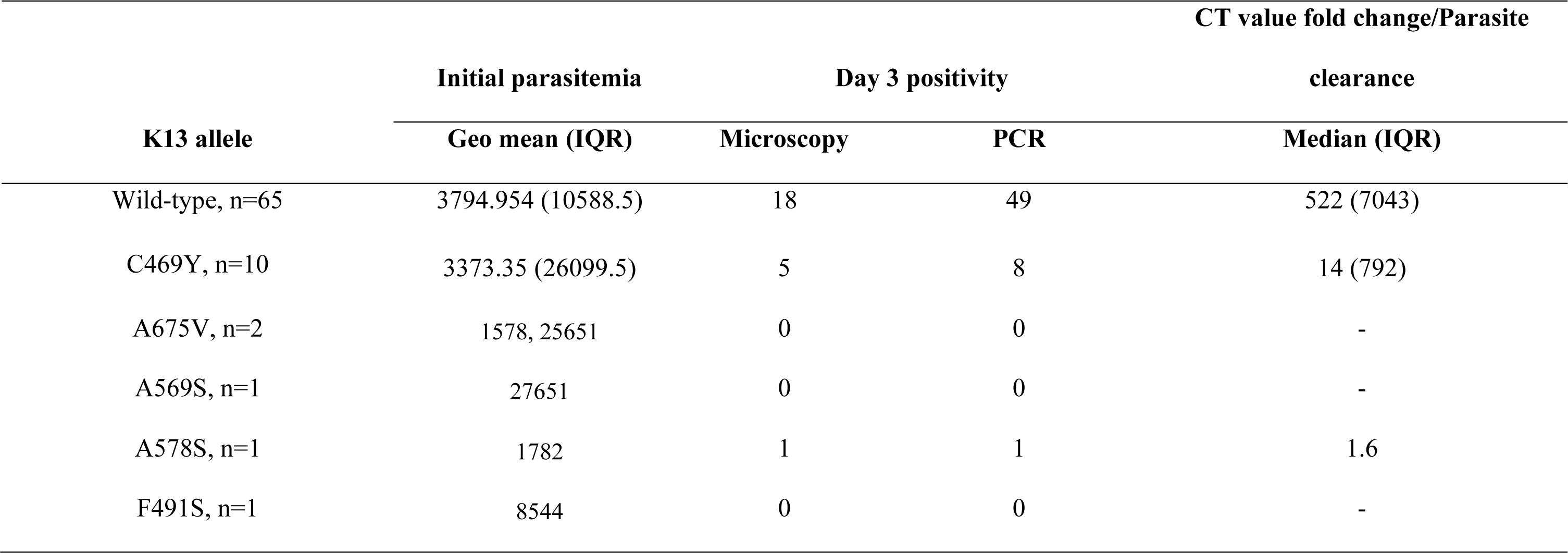
Day 3 positivity and CT value fold change in the K13 alleles from the parasites sequenced.

## Discussion

Artemether-lumefantrine (AL), the first-line treatment for uncomplicated malaria in Uganda, at the current standard fixed weight-based dose regimen, has been pivotal in reducing malaria-related morbidity and mortality. However, emerging *P. falciparum K13* mutations associated with reduced *P. falciparum* susceptibility to AL threaten malaria control and elimination efforts in sub-Saharan Africa. Reduced susceptibility to AL that may be characterised by the persistence of the parasite on day 3 could be associated with partial artemisinin resistance.

Our study reports that, on day 3, there was a prevalence of 24% *P. falciparum* by microscopy among the patients treated with AL in Adjumani district, Northern Uganda. This could indicate the emergence of partial artemisinin resistance. We observed a high prevalence of *P. falciparum* parasites by microscopy, 24% and quantitative polymerase chain reaction (qPCR), 63%, after three days of AL treatment. This observation aligns with previous studies that have reported similar persistence of parasites beyond the standard three-day treatment period in Africa and elsewhere (30–33). Equally, our qPCR results indicated a prevalence of 63%, consistent with a study by Tadele et al. (28) that reported a 60% PCR prevalence of day three *P. falciparum* parasites among patients treated with AL, which may not be sufficient to indicate the development of partial artemisinin resistance. However, the 24% prevalence by microscopy could be a pointer to the potential emergence of partial *P. falciparum* resistance against AL, the current cornerstone of malaria treatment. WHO advises that a day-three parasitemia in patients on ACT treatment calls for routine monitoring to identify suspected artemisinin partial resistance in *P. falciparum* (34).

The persistence of *P. falciparum* parasites on day 3 raises a concern regarding the current effectiveness of artemether and the efficacy of the AL regimen. Our study found a 24% microscopy positivity for *P. falciparum* on day three, which could indicate a possible emergence of partial artemisinin resistance (35,36). A similar finding on the persistence of *P. falciparum* has been reported previously in AL-treated malaria patients in Uganda (30,37–39). Whether this is attributed to the development of resistance, especially the K13 mutation, or lack of adherence to AL has not been fully established. However, a previous study in Uganda has demonstrated moderate to high adherence to AL in treating uncomplicated malaria, whether taken under supervision or not (40). Day 3 positivity and residual *P. falciparum* are likely to cause an increase in malaria recurrence and transmission to female Anopheles mosquitoes (41), driving infection to people and jeopardising the gain in malaria control.

The study identified nonsynonymous mutations at codons 469, 675, 569, 578 and 491 of the K13 propeller gene. This mutation pattern is consistent with recent findings in Uganda (20,39,42–44). The codons 469 and 675 mutations have been associated with artemisinin resistance in northern Uganda (19,21). This is consistent with our finding that most patients with C469Y-mutated *P. falciparum* had parasites that persisted after three days of AL treatment. Among the K13 mutations identified, patients carrying parasites with C469Y (5/10) and A578S (1/1) mutations remained microscopy-positive on day 3; similarly, qPCR results showed high day 3 positivity (8/10) for the C469Y. Unlike a study by Stoke et al. (45) that did not link the C469Y mutation with parasite resistance or delay in parasite clearance, in this study, C469Y was associated with low parasite fold reduction 14, 792 (median, IQR), compared to the wild-type, an indication of reduced *P. falciparum* clearance. This is consistent with the findings of Balikagala et al. and Tumwebaze et al. (19,21) that demonstrated delayed parasite clearance and resistance. A similar study in Pakistan reported a high association between C469Y and artemisinin resistance (46). Among the patients with *P. falciparum* that remained positive on day 3, we identified one patient (1/1) with A578S mutated parasite having a low parasite fold reduction of 1.6. The A578S mutation has been sporadically reported in sub-Saharan African countries (20,47–49), and a study in Uganda reported some association between A578S and slow parasite clearance (50), contrary to studies elsewhere that could not associate it with artemisinin resistance (51,52).

The prevalence of mutation at codon 675 in the malaria patients was 2.5%, and none of the patients with parasites having the A675V mutation remained *P. falciparum* positive on day 3. Although the A675V mutation was previously associated with resistance in Northern Uganda (19), our current study could not link the few to the persistence of *P. falciparum* on day 3, possibly due to the low prevalence and likelihood of chance finding that they are not positive on day 3. Although the A675V mutation is a candidate mutation for artemisinin resistance, a study by Tumwebaze et al. and Stoke et al. (21,45) reported high parasite clearance among parasites with the A675V mutations. We also identified a mutation at codon position 491, which had previously not been identified anywhere. This finding shows the existence of a reservoir for K13 propeller gene polymorphisms, not only in Uganda but also globally in malaria-endemic places (53,54). The diversity of K13 propeller gene mutations across regions further emphasises the need for continued surveillance and research to understand their implications in artemisinin resistance.

Our study had some limitations. Firstly, parasites detected on day 3 were not confirmed whether they were the same parasite on day 0 or a newly acquired infection. Secondly, we did not confirm adherence to treatment among malaria patients who participated in the study. Third, the parasite load on day 0 was not the same across all the patients; low and high *P. falciparum* infections were all considered; the study considered *P. falciparum* positive patients with no specific parasite load and as long as they were prescribed AL. Fourth, the study used two data points of day 0 and day 3 to determine the parasite clearance through fold changes; this makes the data less likely to precisely establish the linear correlation of the parasite clearance slope and unable to capture the fold change pattern of those who are negative on day 3. Lastly, the clinical outcome of the participants was not evaluated according to the WHO 28- day recommendation for an efficacy study. However, the limitations were unlikely to significantly affect the study’s outcome in determining artemisinin effectiveness and identifying markers of artemisinin resistance. Future studies should consider using multiple data points when using the CT value method to assess parasite clearance in effectiveness and efficacy studies of artemisinin.

## Conclusion

The day 3 prevalence of 24% *P. falciparum* by microscopy in the AL-treated patients could indicate the presence of partial resistance to artemisinin. There is an increase in the prevalence of K13 mutation associated with resistance, especially the C469Y, which is associated with a low-fold change in parasite reduction. There is a need to re-evaluate the effectiveness of artemisinin agents in malaria treatment and ensure continuous monitoring of their efficacy in Uganda.

## Data Availability

All data produced in the present work are contained in the manuscript

## Acknowledgement

We acknowledge the research participants from Adjumani Hospital, the staff of Adjumani Hospital: Mr Amosu Emmanuel (Laboratory Technologist), Mr Iranya Vincent (Clinical Officer), Mr Amaruma Allan Levi (Clinical Officer), and Mr Anyanzo Ben (Community Health Worker). Colleagues at Pharmacology/Research Center for Tropical Disease and Vector Control (RTC) laboratory and Central Diagnostic Laboratory.

## Authors Contributions

Conceived and designed the study: MKA MO NM SLN. Performed the experiments: MKA DS MAT. Analysed the data: MKA MO NM. Contributed reagents/materials/analysis tools: MKA MO VP. Initial drafting of manuscript: MKA. Contributed to the final writing of the manuscript: MKA MO NM SLN DS OD VP MAT.

